# Temporally-Informed Random Forests for Suicide Risk Prediction

**DOI:** 10.1101/2021.06.01.21258179

**Authors:** Ilkin Bayramli, Victor Castro, Yuval Barak-Corren, Emily M. Madsen, Matthew K. Nock, Jordan W. Smoller, Ben Y. Reis

**Affiliations:** Predictive Medicine Group, Computational Health Informatics Program, Boston Children’s Hospital, Boston, MA; Harvard University, Cambridge, MA; Mass General Brigham Research Information Science and Computing, Boston, Massachusetts; Psychiatric and Neurodevelopmental Genetics Unit, Center for Genomic Medicine, Massachusetts General Hospital, Boston, Massachusetts, Center for Precision Psychiatry, Department of Psychiatry, Massachusetts General Hospital, Boston, MA; Department of Psychology, Harvard University, Cambridge, MA; Mental Health Research Program, Franciscan Children’s, Brighton, MA; Department of Psychiatry, Massachusetts General Hospital, Boston, MA; Harvard Medical School, Boston, MA

## Abstract

**Background:** Suicide is one of the leading causes of death worldwide, yet clinicians find it difficult to reliably identify individuals at high risk for suicide. Algorithmic approaches for suicide risk detection have been developed in recent years, mostly based on data from electronics health records (EHRs). These models typically do not optimally exploit the valuable temporal information inherent in these longitudinal data.

**Methods:** We propose a temporally enhanced variant of the Random Forest model - Omni-Temporal Balanced Random Forests (OTBRFs) - that incorporates temporal information in every tree within the forest. We develop and validate this model using longitudinal EHRs and clinician notes from the Mass General Brigham Health System recorded between 1998 and 2018, and compare its performance to a baseline Naive Bayes Classifier and two standard versions of Balanced Random Forests.

**Results:** Temporal variables were found to be associated with suicide risk. RF models were more accurate than Naive Bayesian classifiers at predicting suicide risk in advance (AUC=0.824 vs. 0.754 respectively). The OT-BRF model performed best among all RF approaches (0.339 sensitivity at 95% specificity), compared to 0.290 and 0.286 for the other two RF models. Temporal variables were assigned high importance by the models that incorporated them.

**Discussion:** We demonstrate that temporal variables have an important role to play in suicide risk detection, and that requiring their inclusion in all random forest trees leads to increased predictive performance. Integrating temporal information into risk prediction models helps the models interpret patient data in temporal context, improving predictive performance.

## Introduction

Suicide is the tenth leading cause of death in North America and a leading cause of mortality among people aged 15-24 worldwide.^1^ Every year, 800,000 deaths worldwide are due to suicide,^2^ and suicide-related mortality rates are increasing.^3^ Early and accurate identification of individuals with high suicide risk is critical for the development and deployment of effective suicide prevention strategies. However, predicting suicide risk remains a difficult challenge. A study showing that clinicians’ intuition for predicting suicide is no better than random chance highlights the need for exploring algorithmic approaches to this challenge.^4^

In recent years, the availability of high-dimensional electronic health record (EHR) data has sparked a number of efforts to leverage such data for predicting the risk of suicide and suicide attempts.^5 6 7^ In our previous work, a Naive Bayesian Classifier model based on data from over 1.7 million patients in a large healthcare system (Mass General Brigham) achieved an area under the receiver operating curve (AUC) of 0.77, detecting 45% of suicide attempts an average of 3 to 4 years in advance at a specificity of 90%.^5^ This approach was further validated in five independent healthcare systems.^8^ Because algorithmic methods can incorporate vastly more data than a clinician can integrate within the limitations of a typical clinical encounter, they have been shown to outperform risk prediction by clinicians.^5^ However, whereas clinicians may naturally consider the temporal evolution of patient histories, many common classes of predictive models have mostly been unable to explicitly model time-varying features.

To improve upon these efforts, in the present study we consider more advanced modelling approaches and adapt them to more explicitly handle temporal information. A *Random forest model* (RF) is an ensemble of uncorrelated decision trees built from a random subset of features. Random forests are a powerful class of models with great capacity for learning the joint distribution of features without overfitting. RFs have been used successfully for many prediction tasks^9^ including prediction of sudden cardiac arrest,^10^ depression, and other diseases.^11^ Despite their formidable predictive power, standard RFs are not well-suited for handling certain kinds of temporal and sequential data due to their inability to easily capture trends or extrapolate aggregate patterns.^12^

Adapting random forests to effectively capture temporal risk factors for suicide and other prediction tasks requires improvements to the standard RF approach. In standard RFs, a subset of features is selected at random for inclusion in each tree in order to reduce the correlation between trees, prevent overfitting, and lower the variance of the derived estimator. However, this approach may limit the model’s ability to handle longitudinal EHR data. Feature counts are cumulative over time, and the scale of feature counts increases as the patient accumulates more visits. For example, the number of expected diagnoses in a healthy subject over a two year period is likely to be different than the expected number over a ten year period. Interpreting features that vary over time requires a knowledge of the appropriate temporal scaling. In order to properly interpret such features and determine the correct thresholds for node splits within each constituent tree of a RF, it is important to have access to contextualizing summary temporal information. The standard random forest algorithm selects such summary temporal variables in only some of the trees due to random feature sampling.

We propose an optimization of random forests, called *Omni-Temporal Balanced Random Forests* (OT-BRFs). OT-BRFs differ from standard RFs in that they require the inclusion of temporal variables in each constituent tree of the RF, rather than including temporal variables in only some of the trees through random sampling. Requiring the incorporation of temporal features in each tree ensures that longitudinal effects can be appropriately modeled throughout (Figure 1).

**Figure 1.**
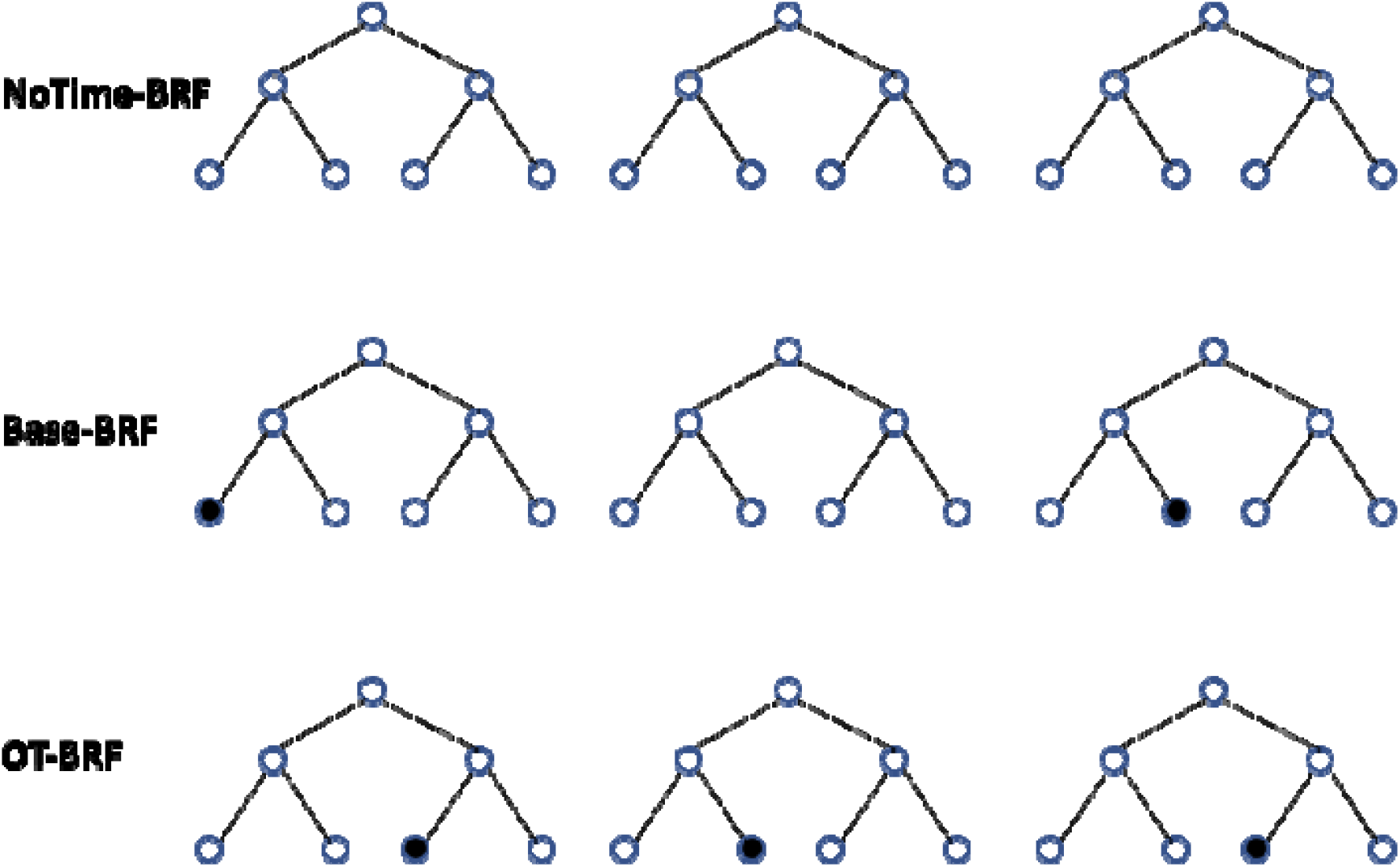
Schematic comparison of Random Forest model types. Three sample trees are shown for each type of RF model. Each tree contains seven nodes. Black nodes represent temporal variables, white nodes represent non-temporal variables. NoTime-BRFs do not include temporal variables. Base-BRFs randomly include temporal variables in some of the trees. OT-BRFs include all temporal variables in each tree.

In the present study, we investigate the performance of both standard RF models and OT-BRFs for suicide prediction, and compare their performance to a baseline Naive Bayes Classifier model. We also examine the top features selected by each model in order to characterize the relative importance of the temporal variables.

## Methods

We analyzed data from the Mass General Brigham Health System’s (MGB) Research Patient Data Registry (RPDR)^13^ - an EHR data warehouse covering 4.6 million patients from two large academic medical centers in Boston (Massachusetts General Hospital and Brigham and Women’s Hospital), as well as additional affiliated community and specialty hospitals in the Boston area. The RPDR was queried for all inpatient and outpatient visits occurring between 1998 and 2018 who met the following inclusion criteria: three or more visits in the individual’s longitudinal record, 30 days or more between the first and last visits, and a least one visit after age 10 and before age 90. For each patient, all demographic, diagnostic, procedure, laboratory, and medication data recorded at each visit were included. We augmented EHR records for each patient with natural language processing (NLP) concept codes extracted from the unstructured clinician notes recorded at each visit.

### Natural Language Processing of clinician notes

In order to generate a set of structured features from the unstructured clinician notes, we created a custom lexicon of suicide-relevant and psychiatric concepts using a variety of approaches including, (1) selecting signs and symptoms and mental and behavioral process semantic types from the Unified Medical Language System (UMLS),^14^ (2) mapping of DSM symptoms and concepts from structured instruments,^15^ (3) applying automated feature extraction from public sources including Wikipedia and MedScape, (4) incorporating Research Domain Criteria (RDoC) domain matrix terms,^15^ (5) selecting predictive features from coded suicide attempt prediction models^16^ and (6) manually annotating terms by expert clinical reviewers. This lexicon was linked to UMLS concepts and included 480 distinct semantic concepts and 1,273 tokens or phrases. Using this lexicon, we ran the HiTex^17^ NLP named-entity extraction pipeline to identify concepts in over 120 million clinical notes. For each note, we identified the presence of a concept (e.g. symptom, disease, mental process) and further tagged concepts as negated (NEG), family history mention (FH) or negated family history (NFH). Negation and family history pipeline components utilized the ConText algorithm.^18^

### Case Definition

Our previous work details the development of the EHR-based case-definition for suicide, also used in this study.^5^ With the assistance of three expert clinicians, we identified codes from *International Classification of Diseases, Ninth Revision* (ICD-9) and *International Classification of Diseases, Tenth Revision* (ICD-10) that captured suicide attempts with a positive predictive value (PPV) greater than 0.70. Individuals having one or more of these codes in their medical record were labeled as a case subject.

We removed all data recorded after the first suicide attempt (index event) for all cases. For the purpose of this study, the case definition was based only on coded diagnostic information in the EHR, and did not include information from the physician’s notes in order to classify individuals as cases vs. non-cases.

### Baseline Model: Naive Bayes Classifier

We used the Naive Bayes Classifier (NBC) model developed in our prior work as a baseline for comparison.^5 8 19^ NBCs are a subclass of Bayesian models that assume strong conditional independence among all input features, reducing model complexity. Highly scalable and interpretable, NBCs have been widely used for clinical decision support tasks.^19^ During model training, NBCs compute a risk score for each feature using the odds ratios of the feature’s prevalence in case and non-case populations, without taking into account interactions between different features. During testing, the NBC risk scores for each concept in a subject’s visit history are added together to generate a summary suicide risk score for the subject.

In this study, each predictor was counted multiple times if it appeared in multiple visits for a given individual. We split our dataset into training and testing sets following a 70/30 split ratio and carried out training using R version 3.6.0 and packages *pROC* and *tidyverse*.

### Random Forest Classifiers

We trained RF models on this same dataset. We used Balanced Random Forest Classifiers ^20^ (BRFs), an extension of Random Forests^21^ designed for label-imbalanced datasets such as the data in this study, where approximately 1 in 100 subjects is a case. BRFs work by either upsampling the minority class, downsampling the majority class, or bootstrap resampling (with replacement) both classes until a specified ratio of classes is met. For our models, we undersampled the majority class to achieve a 1:4 case-control ratio in bootstrap training samples, which we found to be optimal during hyperparameter search.

In representing a subject’s longitudinal record, each row of the subject’s record included the subject’s cumulative visit history (total counts for each feature) up to and including the given visit for that subject. Since an average subject has more than 50 visits in his or her medical history, this data transformation would result in terabytes-large tables with which training a model in a reasonable amount of time would be impractical. Therefore, we sampled from a set of 90,000 subjects for each of the testing and training sets, and included a random sample of visits for each subject (each visit was included with a probability of 0.05). During the sampling of training data, we ensured that the proportion of cases was elevated from 1% to around 12% to help support the bootstrap sampling: Raising the case prevalence to 12% ensures that for each tree, the bootstrap sets resampled from this training data are sufficiently large after random undersampling of the majority class to a 1:4 case-control ratio. The test set maintained the natural 1% suicide prevalence.

We performed variable selection as follows: We kept EHR features (diagnoses, labs, medications) that occurred in at least 10 non-case and 10 case subjects. We also examined the risk scores derived during the fitting of the aforementioned NBC model, and eliminated all features with an absolute risk score of less than 0.1. Since the number of NLP features was very small (N=2,373) relative to the EHR data types (N = 43,435), we did not eliminate any NLP features.

We included three key summary temporal variables: Period covered - the time in years since the patient’s first visit; *Visit number* - the count of visits since the patient’s first visit; and *Visit rate* -the period covered divided by number of visits.

We examined three variations of the BRF model: 1) The NoTime-BRF model was trained on the EHR and NLP data, without the three temporal summary variables defined above; 2) The Base BRF model was trained on EHR, NLP and the three temporal summary variables, with temporal variables treated similarly to other variables and selected randomly to be included in some of the trees in the forest; and 3) The *Omni-Temporal Balanced Random Forest Classifier* (OT-BRF) model - this model requires each tree in the forest to include all three temporal variables, while randomly sampling the rest of the EHR and NLP features.

For hyperparameter optimization, we performed a grid search with 5-fold cross-validation on the BRF parameter space. This yielded a model with 30 trees, 50% of all features sampled for each tree, size of bootstrap samples set to the total number of samples, and 1:4 case-control ratio in every bootstrap sample by undersampling of the majority class. We used Python version 3.6.9 with the libraries *scikit-learn, imblearn, numpy*, and *pandas*.

### Feature Importance

To evaluate the relative importance of various features in the random forest models, we used the impurity-based Gini importance metric. We computed the normalized Gini importance for a given feature as the total decrease in node impurity resulting from inclusion of that feature, averaged across all trees.

### Ethics

This research was approved by the Massachusetts General Hospital Institutional Review Board, along with an IRB reliance agreement from the Boston Children’s Hospital Institutional Review Board.

## Results

NBC models were trained on data from 1,625,350 patients, consisting of 1,608,806 non-cases (99%) and 16,544 cases (1.0%). The testing set included a total of 697,411 patients with 7,155 of those being cases (1.0%). The NBC models used all 45,808 EHR and NLP features available in the dataset.

For developing the RF models, the sampled dataset contained 231,034 unique visits of 52,006 patients in the training set and 226,868 visits of 52,664 patients in the testing set. Cases constituted 19.4% and 0.9% of training and testing patients, respectively. Applying the feature selection steps described above yielded 16,131 features used in RF model training and testing. Figure 2 shows that the distribution of visit numbers in this dataset was biased towards relatively small counts: patients with 5, 10, 20 or more visits by definition had data for their 1st, 2nd, 3rd visits as well, so earlier visit counts were likely to be sampled more frequently.

**Figure 2.**
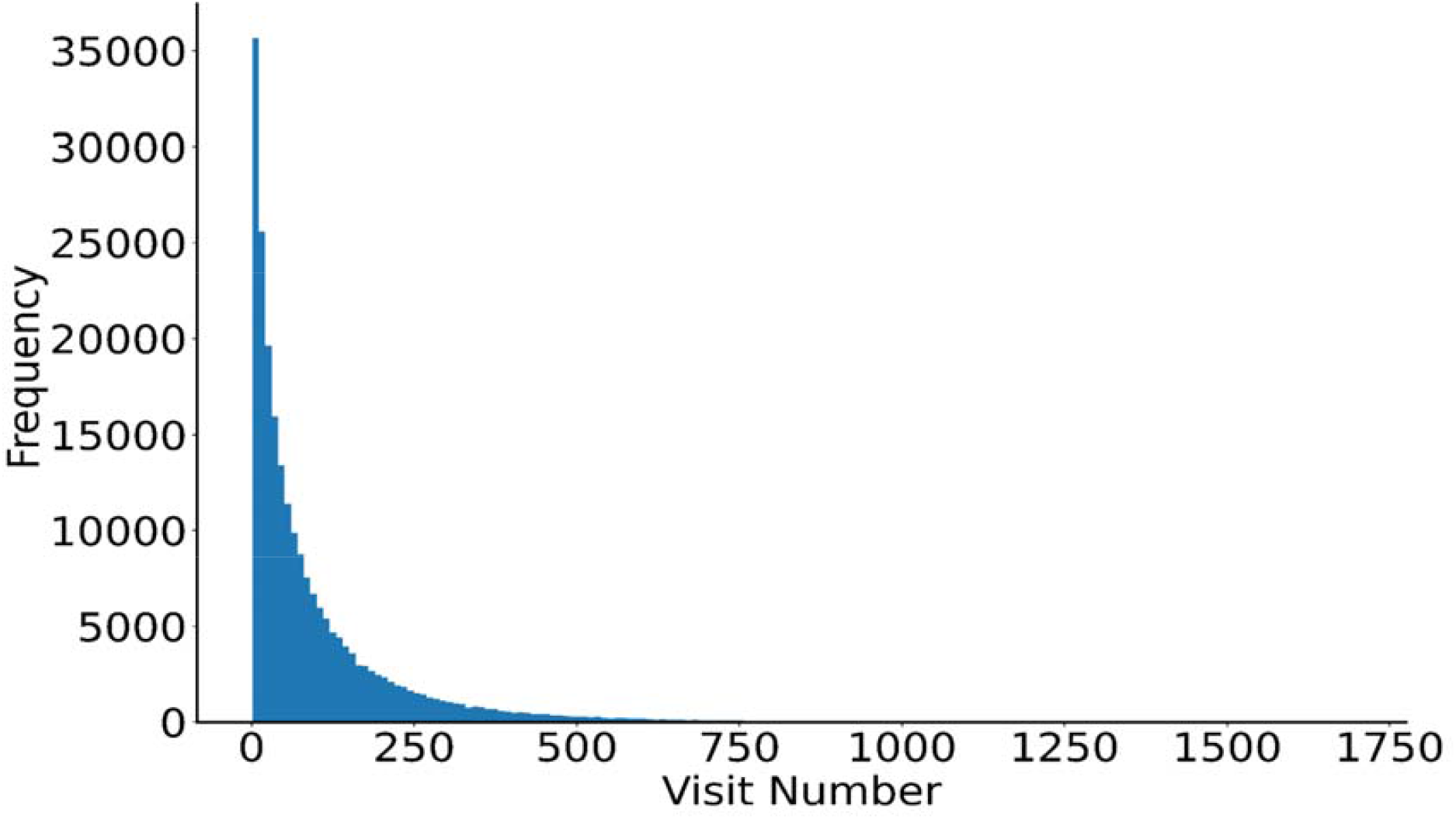
Distribution of visit numbers in the test set. Most individuals had few visits in their EHR, while some had many visits.

### Univariate Analyses of Temporal Variables

Figure 3 shows the proportion of suicide cases plotted against different ranges of values for each of the three temporal variables. Figure 3a shows that suicide attempt rates are slightly below average for small visit numbers (1-10), approximately average for moderate and large visit numbers (10 - 350), and significantly above average for very high visit numbers (350+), consistent with greater opportunities for observation. Figure 3b shows below-average suicide attempt rates for patients with short (<1 year) or long (>14 years) coverage periods, and above-average for the intermediate range of 1-14 years coverage. Figure 3c shows elevated suicide risk among patients with fewer than 0.5 visits per year, and generally lower risk for higher visit rates.

**Figure 3.**
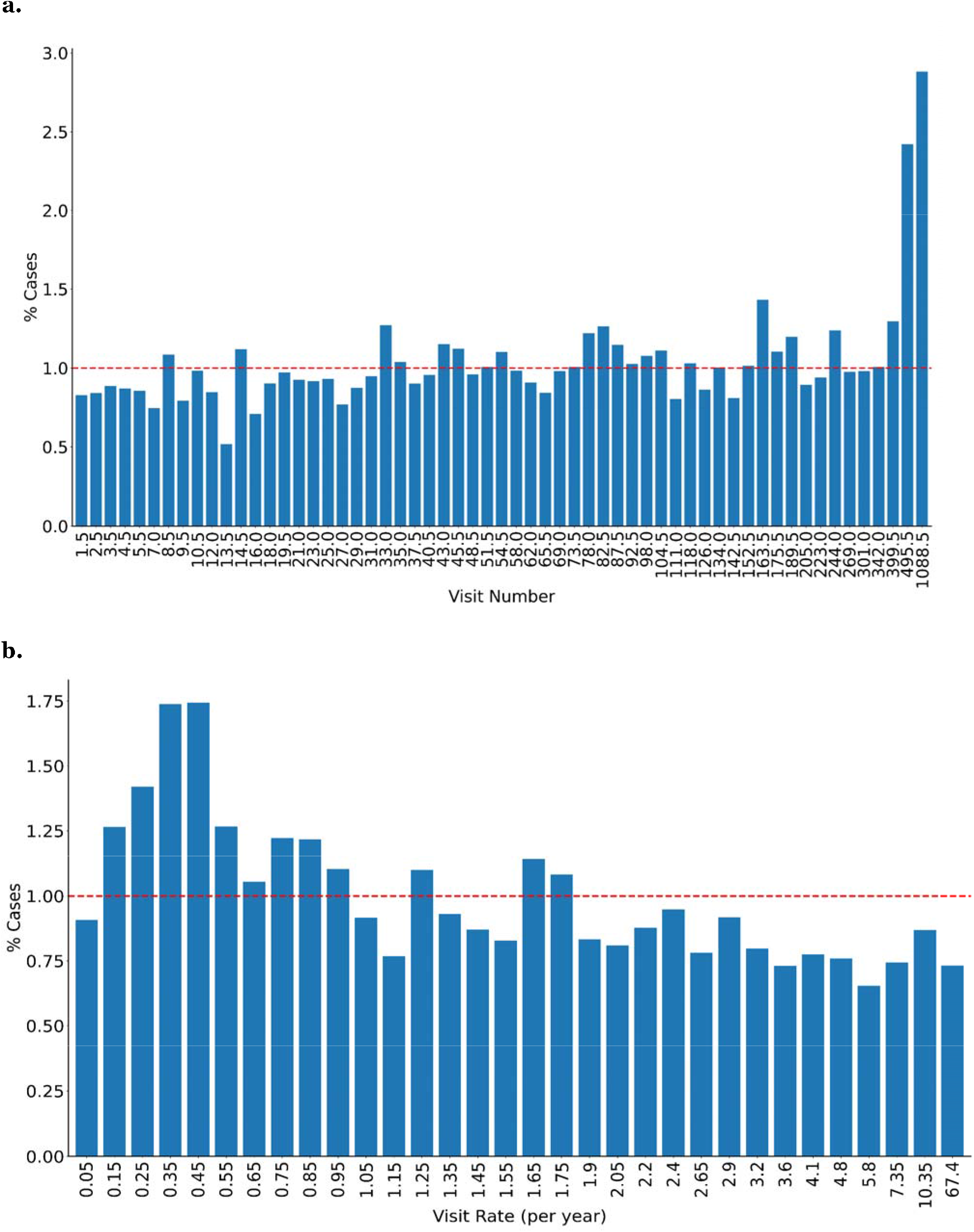
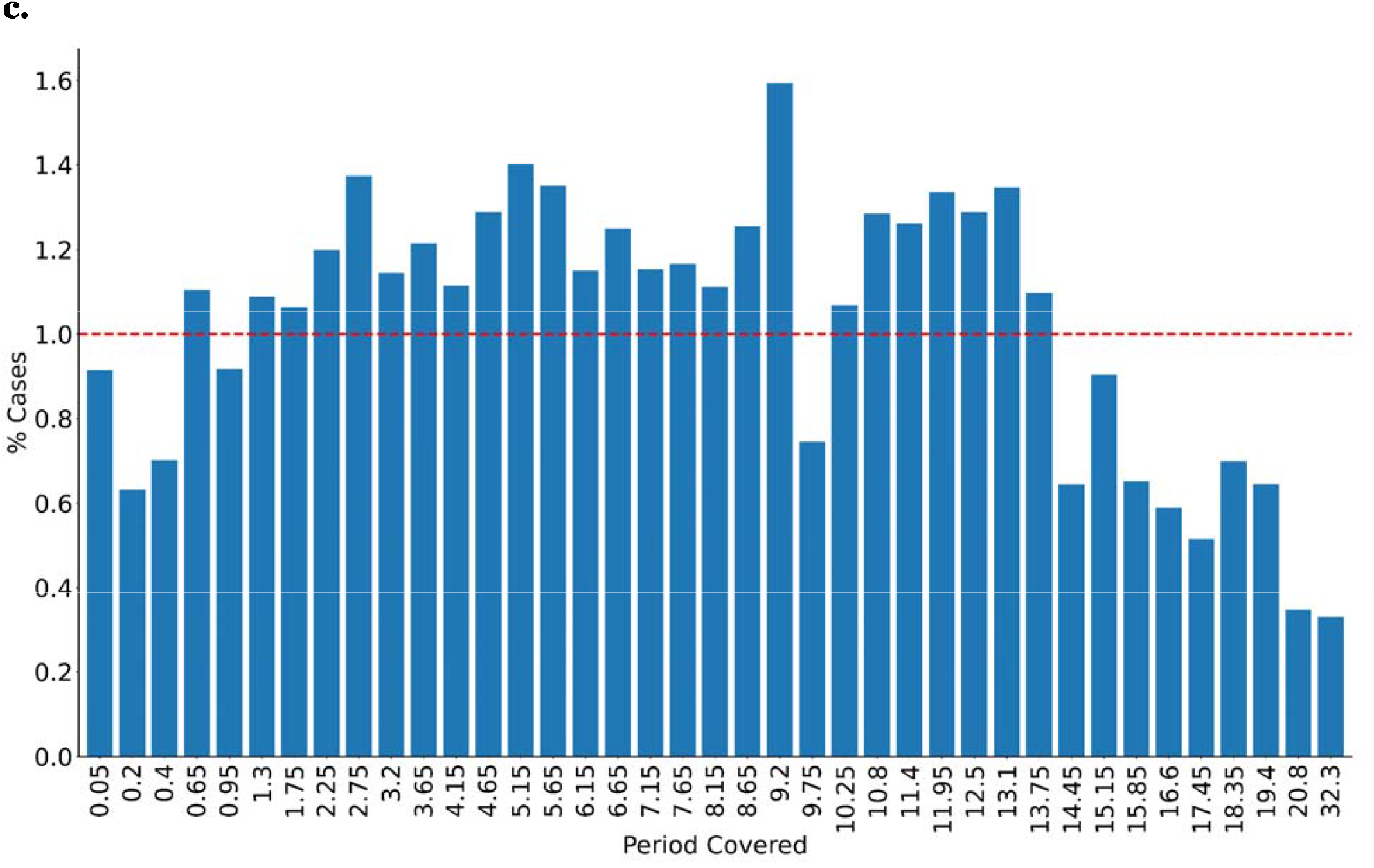
Bivariate relationships between temporal variables and suicide risk. **a**. Proportion of suicide cases for different total-visit-number ranges, **b**. visit-rate ranges, and **c**. total-period-covered ranges.

### Model Comparisons

Model performance metrics for the NBC and BRF models are presented in Table 1. All three RF models performed better than the baseline NBC model in terms of AUC, PPV and sensitivity across all specificity thresholds examined. Among RF models, for all specificity thresholds, the OT-BRF model performed better than Base-BRF, which in turn performed better than NoTime-BRF. At 95% specificity, OT-BRF outperformed Base-BRF by 0.6% in PPV (relative improvement of 9.8%) and 4.9% in sensitivity (relative improvement of 16.9%).

**Table 1.** Model performance comparison. Sensitivity and PPV values for the four tested models at different specificity levels, together with AUC values: NBC - Naive Bayesian Classifier; NoTime-BRF - Balanced Random Forest with no temporal variables; Base BRF - Balanced Random Forest with temporal variables; OT-BRF -Omni-time Balanced Random Forest with required inclusion of temporal variables in each tree. Boldface indicates highest values in each row.

|  | NBC |  | NoTime-BRF |  | Base BRF |  | OT-BRF |  |
| --- | --- | --- | --- | --- | --- | --- | --- | --- |
| Specificicity | PPV | Sensitivity | PPV | Sensitivity | PPV | Sensitivity | PPV | Sensitivity |
| **0.80** | 0.032 | 0.550 | 0.032 | 0.640 | 0.033 | 0.649 | **0.034** | **0.656** |
| **0.90** | 0.042 | 0.403 | 0.043 | 0.463 | 0.043 | 0.468 | **0.045** | **0.499** |
| **0.95** | 0.051 | 0.272 | 0.060 | 0.286 | 0.061 | 0.290 | **0.067** | **0.339** |
| **AUC** | 0.754 |  | 0.808 |  | 0.810 |  | **0.824** |  |

We further examined the performance of the RF models with different amounts of patient data available. Table 2 and Figure 4 show that the performance of all three RF models remained relatively stable over different visit count intervals, never falling below 0.77 AUC. For all three RF models, the performance peaked at 39-80 visits. For all patients with data on 15 or more visits, the OT-BRF model performed slightly better than the other two random forest models. For patients with only 1-15 visits, the best-performing model was NoTime-BRF. Since most patients in this group have only 1-3 visits for which temporal variables are ill-defined or very noisy, the slightly poorer performance of models that use temporal variables in this interval is to be expected.

**Table 2.** Testing-set-AUCs of the three random forest models for patients with different total-visit-number ranges. Boldface indicates highest values in each column.

| Visit Number | 1 - 14 | 15 - 39 | 40 - 79 | 80 - 165 | 166 - 3297 | All Visits |
| --- | --- | --- | --- | --- | --- | --- |
| NoTime-BRF | **0.791** | 0.821 | 0.821 | 0.821 | 0.779 | 0.808 |
| BRF | 0.788 | 0.828 | 0.837 | 0.808 | 0.775 | 0.810 |
| OT-BRF | 0.784 | **0.844** | **0.843** | **0.825** | **0.809** | **0.824** |

**Figure 4.**
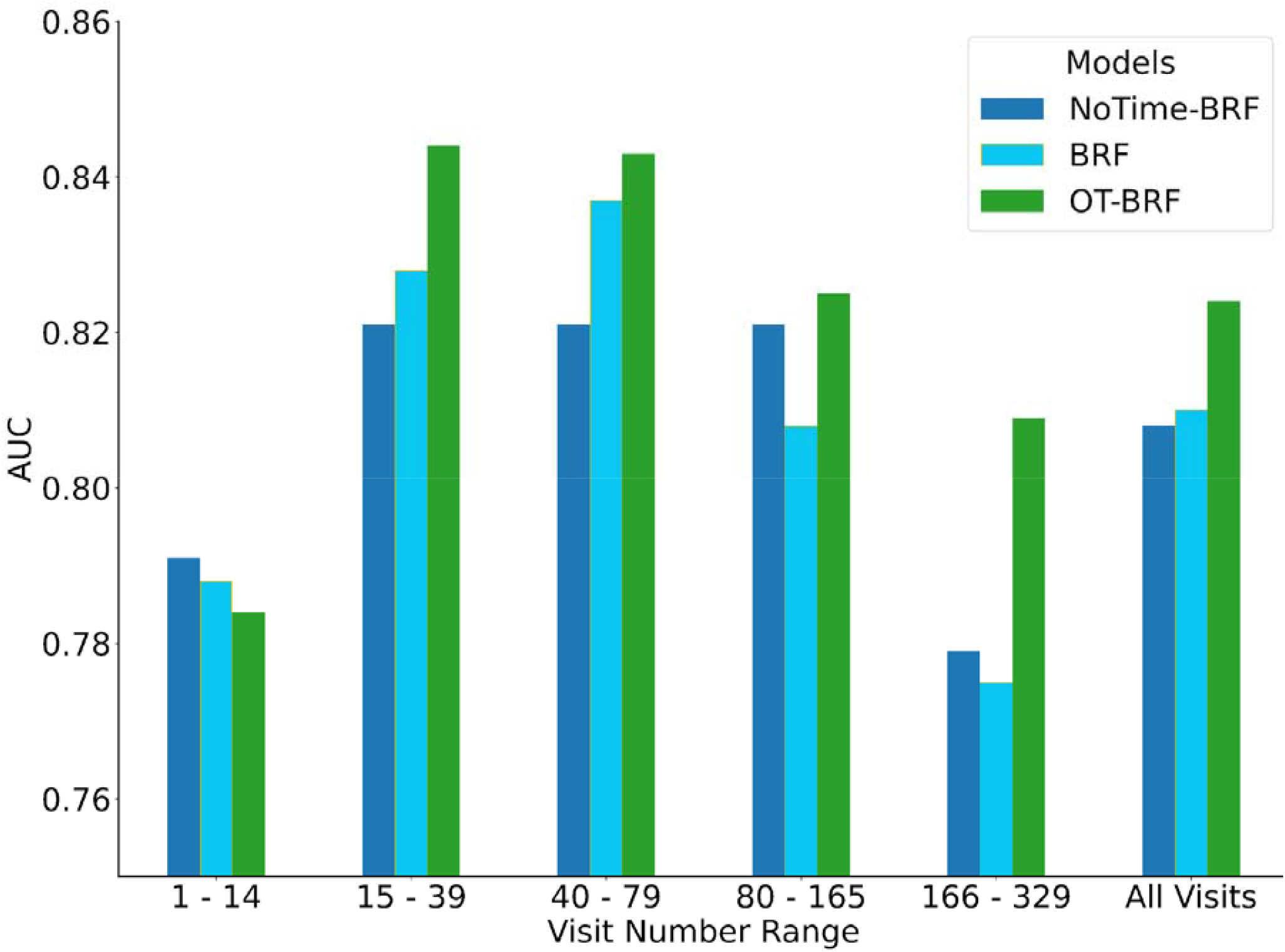
Model performance comparison. Area under the receiver operating curve (AUC) of the three models as a function of the number of visits per subject.

### Feature Importance

We compared the feature importances of the Base-BRF and OT-BRF models to see how the required inclusion of temporal variables in all trees of the Random Forest affects their contribution to model performance. Table 3 presents the top 20 normalized feature importances of OT-BRF with their corresponding ranks in Base-BRF. In both models, the three temporal variables were ranked very high - in the top 20 from among 16,131 included features. However, these variables were assigned greater importance in OT-BRF than in Base-BRF: the variables “period covered”, “visit number”, and “visit rate” have ranks 3, 10 and 11 respectively in OT-BRF, compared with ranks 8, 14, and 11 respectively in Base-BRF.

**Table 3.** Top 20 features, ranked by feature importance. Feature importance calculated using Gini mean decrease in impurity. Features are shown sorted by OT-BRF importances in descending order. (EHR = concept from structured electronic health record; NLP = concept from unstructured clinician notes analyzed with natural language processing techniques; DEMO = Demographic variables; TEMP = Temporal variables)

| Concept Name | Data Type | Import. OT-BRF | Import. Base-BRF | Rank OT-BRF | Rank Base-BRF |
| --- | --- | --- | --- | --- | --- |
| Drug screen, qualitative; multiple drug classes | EHR | 0.03712 | 0.04634 | 1 | 1 |
| Impulse-control disorder | NLP | 0.02733 | 0.03217 | 2 | 2 |
| Period covered | TEMP | 0.02118 | 0.01330 | 3 | 8 |
| Chloride; blood | NLP | 0.01815 | 0.01091 | 4 | 12 |
| Age | DEMO | 0.01622 | 0.02221 | 5 | 4 |
| Recreational drug use | NLP | 0.01486 | 0.01315 | 6 | 9 |
| Marital Status - married | DEMO | 0.01466 | 0.02234 | 7 | 3 |
| Current zip median income | DEMO | 0.01446 | 0.02048 | 8 | 6 |
| Estab Pt - Low Level Visit HC | EHR | 0.01414 | 0.02064 | 9 | 5 |
| Visit number | TEMP | 0.01306 | 0.00858 | 10 | 14 |
| Visit rate | TEMP | 0.00996 | 0.01093 | 11 | 11 |
| Depressive disorder, not elsewhere classified | EHR | 0.00982 | 0.01559 | 12 | 7 |
| Suicide thoughts | NLP | 0.00958 | 0.00669 | 13 | 15 |
| Psychiatric diagnostic interview examination | EHR | 0.00752 | 0.00580 | 14 | 17 |
| Encounter, general adult medical examination | EHR | 0.00611 | 0.00126 | 15 | 71 |
| Emergency department visit for the evaluation | EHR | 0.00544 | 0.00543 | 16 | 18 |
| Pharmacologic management, incl. prescription. | EHR | 0.00463 | 0.00586 | 17 | 16 |
| Suicide attempts | NLP | 0.00436 | 0.00390 | 18 | 21 |
| Collection Venous Blood Venipuncture HC | EHR | 0.00420 | 0.01198 | 19 | 10 |
| Individual psychotherapy, insight oriented, behavior | EHR | 0.00405 | 0.00468 | 20 | 19 |

Examining some of the 30 trees built by the OT-BRF we found temporal variables were chosen for inclusion at both relatively high-level positions near the root node of the tree, as well as relatively low-level positions near the leaf nodes.

## Discussion

The results of this study show that incorporating temporal variables into predictive models of suicide risk improved performance. The predictive improvement resulting from the inclusion of temporal variables was observed across a range of longitudinal history lengths. RF models performed better than NBC models, even though the RF models were trained using only a subsample of visits for a subsample of patients, while the NBC model was trained on all visits from all patients in the study. This is likely due to the ability of RF models to capture more complex relationships, including interactions between features. The Balanced RF modeling framework allows for reducing the imbalance in the training dataset, further improving performance.

Comparing the performance of the different RF models, those that included temporal variables in some of their trees (Base BRFs) performed better than RF models that did not include any temporal variables whatsoever (no-time BRFs). RF models that required inclusion of all the temporal variables in each of their trees (OT-BRF) yielded the best performance. Our analysis of the impurity-based feature importances and the trained decision trees was consistent with the OT-BRFs making better use of temporal variables than the baseline RFs.

Temporal variables were associated with suicide risk. We found elevated suicide risk among patients with fewer than 0.5 visits per year, and generally lower risk for those with higher visit rates. This may be because people with few visits are isolating themselves from the healthcare system, which may be associated with general isolation and increased suicide risk. It may also be due to the fact that individuals who have more frequent and regular contact with a healthcare provider can be monitored for psychiatric deterioration in advance and supported with various measures to prevent future suicide attempts.

The inclusion of temporal variables at high-level nodes of RF trees confirms the importance of the time-related features. Their inclusion at the bottom nodes of the tree - sometimes multiple times back to back - right before the final classification decision is made, is consistent with the scale-adjusting function of temporal variables described above.

For patients having at least 15 visits in their longitudinal record (and thus well-defined temporal summary variables), OT-BRFs consistently outperformed all other models tested in terms of AUC and PPV. However, in some respects the improved performance of OT-BRFs was modest: Not including temporal variables in any tree (NoTime-BRFs) resulted in a 1.8% drop in validation AUC compared to the OT-BRFs that include temporal variables in all trees. This modest change in performance may be related to the choice of splitting thresholds in the RF trees. Most trees split their nodes at rather small thresholds such as 0.5 or 1.5, even for concepts that frequently occur 6-7 times in individual subjects. In other words, rather than capturing how many times a concept has occurred in a subject, the model captures only a binary fact - whether the concept has occurred at all. Binary data are inherently immune to the scale problem described early on. This may explain the relatively modest drop in the predictive power of trees when they do not use temporal variables at all.

We would expect the boost in performance from including temporal variables to be even greater if more trees captured multiple time-dependent thresholds for the same feature. For example, Figure 3 shows that the EHR concept of “general medical examination” was assigned a rank of 15 by OT-BRF compared to a rank of 71 by the base BRF. For this feature, a binary yes/no flag is less informative since almost all patients in the study have one or more doctor visits. However, the expected number of this feature increases over time. Trees lacking the temporal information to put the scale of the feature into temporal context, could struggle to draw useful insights from it. We believe this may be the reason why OT-BRF ranks this feature as highly influential for predicting suicide, while Base-BRFs rank this same feature far less highly. This is a topic for further research.

This study has a number of limitations. The key mechanism employed by RF models for reducing correlation between trees is the random sampling of features for each tree. Uncorrelated trees reduce the estimator’s variance while increasing its bias, which has been shown to result in higher generalization accuracy. Our proposed approach risks increasing the correlation between trees through uniform inclusion of the same three temporal variables. However, we expect this effect to be quite negligible since the rest of the approximately 16,000 features are still sampled randomly for each tree. Indeed, the net effect observed is an improvement in prediction performance. The impurity-based cost functions described in the methods section have well-known drawbacks, which have been known to lead to high importance being assigned to features that have a different scale or high number of categories, even if these features are unimportant.^22^ However, most variables ranked in the top 20 in feature importance -- such as drug screening, recreational drug use, suicidal ideation, age, depressive disorder, prior suicide attempt, impulse control disorder -- are commonly known to be clinically associated with suicide risk. This validation indicates that our study does not seem to suffer from these potential shortcomings, likely due to the fact that most included features have similar scales and number of categories.

Several researchers have proposed extensions of random forests for temporal datasets. For the task of sensor-based human activity recognition, Ooi et al.^23^ introduced two modifications to the random forest algorithm: 1) a *temporal sampling mechanism* that rearranges the sensor data by considering its sequence of occurrence, and (2) *temporal randomization* that modifies the randomization procedure in RFs to preserve the temporal sequence of captured sensor data. Nitze et al. combined RFs with time-series for multi-temporal land-cover classification.^24^ Gomes et al. proposed *Adaptive Random Forests* for evolving data stream classification.^25^

We introduce a new modeling strategy that modifies RF methods by requiring the inclusion of time-related summary variables throughout the RF. We show that this approach outperforms both our validated NBC models and other RF approaches in the prediction of suicide risk. This approach can be used by researchers and practitioners to better exploit temporal information and improve patient risk prediction.

## Data Availability

The data used in this study cannot be made publicly available due to restrictions relating to the use of electronic health record data.

## Acknowledgements

This work was supported in part by a gift from the Tommy Fuss Fund and R01MH117599 (Drs. Smoller and Reis) from the National Institute of Mental Health.

## Author Contributions

IB and BYR conceived and planned the research, together with input from all other authors. IB conducted the principal data analysis and modeling work. VC conducted the NLP analysis and prepared the datasets for analysis. YBC also contributed to the modeling work. BYR supervised the research. IB, VC, YBC, EM, MKN, JWS and BYR all discussed the results and contributed to the final manuscript.

## References

1. Fazel, S. & Runeson, B. Suicide. N. Engl. J. Med. 382, 266–274 (2020).

2. Naghavi, M. & Global Burden of Disease Self-Harm Collaborators. Global, regional, and national burden of suicide mortality 1990 to 2016: systematic analysis for the Global Burden of Disease Study 2016. BMJ 364, 94 (2019).

3. Hedegaard, H., Curtin, S. C. & Warner, M. Suicide Rates in the United States Continue to Increase. NCHS Data Brief 1–8 (2018).

4. Nock, M. K. et al. Measuring the suicidal mind: implicit cognition predicts suicidal behavior. Psychol. Sci. 21, 511–517 (2010).

5. Barak-Corren, Y. et al. Predicting Suicidal Behavior From Longitudinal Electronic Health Records. Am. J. Psychiatry 174, 154–162 (2017).

6. Simon, G. E. et al. Predicting Suicide Attempts and Suicide Deaths Following Outpatient Visits Using Electronic Health Records. Am. J. Psychiatry 175, 951–960 (2018).

7. Walsh, C. G., Ribeiro, J. D. & Franklin, J. C. Predicting suicide attempts in adolescents with longitudinal clinical data and machine learning. J. Child Psychol. Psychiatry 59, 1261–1270 (2018).

8. Barak-Corren, Y. et al. Validation of an Electronic Health Record–Based Suicide Risk Prediction Modeling Approach Across Multiple Health Care Systems. JAMA Netw Open 3, e201262–e201262 (2020).

9. Wyner, A. J., Olson, M., Bleich, J. & Mease, D. Explaining the success of adaboost and random forests as interpolating classifiers. J. Mach. Learn. Res. 18, 1558–1590 (2017).

10. Wongvibulsin, S., Wu, K. C. & Zeger, S. L. Clinical risk prediction with random forests for survival, longitudinal, and multivariate (RF-SLAM) data analysis. BMC Med. Res. Methodol. 20, 1 (2019).

11. Khalilia, M., Chakraborty, S. & Popescu, M. Predicting disease risks from highly imbalanced data using random forest. BMC Med. Inform. Decis. Mak. 11, 51 (2011).

12. Moon, J., Kim, Y., Son, M. & Hwang, E. Hybrid Short-Term Load Forecasting Scheme Using Random Forest and Multilayer Perceptron. Energies 11, 3283 (2018).

13. Nalichowski, R., Keogh, D., Chueh, H. C. & Murphy, S. N. Calculating the benefits of a Research Patient Data Repository. AMIA Annu. Symp. Proc. 1044 (2006).

14. Bodenreider, O. The Unified Medical Language System (UMLS): integrating biomedical terminology. Nucleic Acids Res. 32, D267–70 (2004).

15. Ross, J. Psychiatric Phenotyping Using Symptom Profiles: Can Self-Report Symptoms Inform a New Psychiatric Taxonomy? (UCSF, 2018).

16. McCoy, T. H., Jr et al. High Throughput Phenotyping for Dimensional Psychopathology in Electronic Health Records. Biol. Psychiatry 83, 997–1004 (2018).

17. Zeng, Q. T. et al. Extracting principal diagnosis, co-morbidity and smoking status for asthma research: evaluation of a natural language processing system. BMC Med. Inform. Decis. Mak. 6, 30 (2006).

18. Chapman, W., Dowling, J. & Chu, D. ConText: An algorithm for identifying contextual features from clinical text. in Biological, translational, and clinical language processing 81–88 (2007).

19. Reis, B. Y., Kohane, I. S. & Mandl, K. D. Longitudinal histories as predictors of future diagnoses of domestic abuse: modelling study. BMJ 339, b3677 (2009).

20. Chen Chao, Andy Liaw, and Leo Breiman. Using random forest to learn imbalanced data. 1–12 (2004).

21. Breiman, L. Random Forests. Mach. Learn. 45, 5–32 (2001).

22. Strobl, C., Boulesteix, A.-L., Zeileis, A. & Hothorn, T. Bias in random forest variable importance measures: illustrations, sources and a solution. BMC Bioinformatics 8, 25 (2007).

23. Ooi, S. Y., Tan, S. C. & Cheah, W. P. Classifying Human Activities with Temporal Extension of Random Forest. in Neural Information Processing 3–10 (Springer International Publishing, 2016).

24. Nitze, I., Barrett, B. & Cawkwell, F. Temporal optimisation of image acquisition for land cover classification with Random Forest and MODIS time-series. Int. J. Appl. Earth Obs. Geoinf. 34, 136–146 (2015).

25. Gomes, H. M. et al. Adaptive random forests for evolving data stream classification. Mach. Learn. 106, 1469–1495 (2017).

